# Pixel-Level Quantification of Subcutaneous Shear-Wave Velocity Heterogeneity for Grading Lymphatic Obstruction in Breast Cancer-Related Lymphedema

**DOI:** 10.64898/2026.09.25.26364003

**Authors:** Zheng-Yu Hoe, Chen-Pin Chou, Yen-Dun Tzeng, Ruei-Sian Ding, Cheng-Tang Pan, Ming-Chan Lee, Chao-Hsien Lee

## Abstract

**Objectives:** To develop and validate pixel data analysis (PDA) for reconstructing pixel-level shear-wave velocity (SWV) from saved Holder-Optimized Elastography (HOE) elastograms and to determine whether whole-field subcutaneous heterogeneity provides a quantitative signature of lymphatic obstruction compared with visual high-velocity-area (HVA) counting and sparse manual region-of-interest (ROI) sampling.

**Materials and Methods:** In this cross-sectional study, 110 women after breast cancer surgery underwent lymphoscintigraphy and HOE, a pressure-minimized acquisition framework that preserves conventionally captured SWV information while additionally revealing spatial abnormalities attenuated by handheld acquisition. Reconstruction validation used 200 paired ROIs, spanning nearly the full displayed SWV range, from 49 images retrieved from the picture archiving and communication system; reproducibility was assessed across 96 matched anatomical sites from 6 participants undergoing independently reacquired HOE examinations. PDA, visual HVA counting, and manual ROI sampling were evaluated on the same HOE-acquired elastograms in task-specific common complete-case cohorts.

**Results:** PDA reconstructed the machine-reported SWV with near-zero error over a range of 1.17 - 9.97 m/s (mean difference, +0.0009 m/s; mean absolute error, 0.0075 m/s; 95% limits of agreement, -0.0202 to +0.0221 m/s). For subcutaneous variance, interobserver/intraobserver intraclass correlation coefficients were 0.709/0.799 for PDA and 0.178/0.560 for manual ROI sampling. In the primary no obstruction versus partial obstruction comparison (n = 93), forearm-pooled PDA subcutaneous variance ratio yielded the highest observed area under the curve (AUC, 0.858; 95% confidence interval, 0.771 – 0.946), compared with 0.820 for visual HVA counting and 0.794 for manual ROI sampling. PDA’s subcutaneous variance ratio remained a leading feature across all three comparisons (AUC, 0.858 – 0.878), including partial versus total obstruction grading, where all three frameworks showed high discrimination.

**Conclusions:** PDA accurately reconstructs pixel-level SWV from saved HOE elastograms and enables reproducible whole-field quantification of superficial shear-wave elastography heterogeneity. Subcutaneous variance ratio was the leading quantitative PDA feature across lymphatic-obstruction comparisons and was particularly informative in partial obstruction. The same approach also enables retrospective quantitative reanalysis of compatible archived elastograms.

**Plain-Language Summary:** Pixel data analysis (PDA) reconstructs quantitative pixel-level shear-wave velocity from saved superficial elastography images, allowing the complete measurable color field to be analyzed rather than a few manually sampled regions. Reconstructed values closely matched machine measurements, and whole-field subcutaneous variance was substantially more reproducible than manual ROI sampling. Subcutaneous variance ratio remained a leading diagnostic signal across obstruction severities, including in partial obstruction without visually detectable high-velocity areas.

## Introduction

Breast cancer-related lymphedema (BCRL) is a common and often persistent complication after breast cancer treatment that can impair upper-limb function, quality of life, and survivorship^1–3^. Lymphoscintigraphy remains the imaging reference standard for identifying lymphatic obstruction and grading its severity^4,5^, but its need for radiotracer injection and its limited practicality for short-interval follow-up constrain routine serial use^6^. A complementary, noninvasive method for repeated evaluation of upper-limb lymphatic impairment therefore remains needed.

Shear-wave elastography (SWE), including acoustic radiation force impulse (ARFI)-based techniques, offers an ultrasound-based means of assessing tissue mechanical changes^7,8^, but prior lymphedema studies have reported variable findings across tissue layers, scan sites, disease stages, and loading conditions^9–15^. Conventional superficial SWE can lose spatial information at two stages: acquisition-related loading can attenuate or distort superficial heterogeneity, and subsequent sparse region-of-interest (ROI) sampling can incompletely represent the remaining spatial distribution. This analytic limitation is particularly relevant in lymphedema, because MRI demonstrates spatially nonuniform and stage-dependent subcutaneous fluid infiltration^16,17^.

To reduce acquisition-related information loss, we developed Holder-Optimized Elastography (HOE), a pressure-minimized framework that stabilizes the transducer and minimizes superficial tissue loading (Supplement 1; S. Figure 1). In the related methodological study, HOE preserved conspicuous subcutaneous high-velocity areas (HVAs) and broader shear-wave velocity (SWV) heterogeneity that were markedly attenuated during conventional handheld acquisition^18^. Correlative MRI, D2-40 histopathology, and intraoperative indocyanine-green observations showed strong spatial correspondence between HVA-localized regions and obstruction-associated lymphatic structures. Visual HVA counting captured conspicuous focal abnormalities but did not quantify the broader heterogeneous field. In partial obstruction, the abnormal pattern may be diffuse and spatially patchy rather than dominated by discrete high-velocity foci. Exploiting the additional spatial information preserved by HOE therefore requires a whole-field quantification method.

However, the system used in the methodological study^18^ did not provide an exportable pixel-level SWV matrix for retrospective analysis. We therefore developed pixel data analysis (PDA) to separate the grayscale and SWV-encoded components of saved composite elastograms, reconstruct physical SWV for each measurable color pixel, and quantify the complete layer-specific distribution. Because the method operates on preserved composite images, it can also be applied retrospectively to compatible archived images when the color/SWV encoding has been maintained through a non-destructive storage pipeline.

The present study developed and validated PDA, compared its whole-field measurements with visual HVA counting and sparse manual ROI sampling performed on the same HOE-acquired elastograms, and evaluated whether subcutaneous heterogeneity could characterize no, partial, and total lymphatic obstruction.

## Materials and Methods

### Study design and cohort relationship

This single-center cross-sectional analysis was approved by the institutional review board of [blinded for peer review] (approval no. [blinded for peer review]), and all participants provided written informed consent before enrollment. Medical history review, clinical evaluation, lymphoscintigraphy, and HOE-based ARFI elastography were completed within a 2-week interval.

The present analysis used prospectively acquired HOE elastograms from the clinical cohort described in the related HOE methodological report^18^. HOE provided the shared pressure-minimized acquisition framework that was used as the basis for all comparisons. The index analysis in this report was offline PDA reconstruction and its subsequent whole-field quantification, and visual HVA counting and manual ROI analysis were applied to the same HOE-acquired elastograms as its comparators.

### Participants and reference standard

Between October 2022 and July 2024, women aged 20 years or older who had undergone breast cancer surgery were recruited from our institution. The exclusion criteria were unhealed wounds or other upper-limb conditions unsuitable for elastography; bilateral upper-limb swelling from causes other than breast cancer-related lymphedema; taxane chemotherapy within 6 months; cellulitis in either upper limb within 6 weeks; unreadable lymphoscintigraphy; or incomplete clinical or imaging assessments. Participant flow, cohort composition, and analysis subsets are summarized in Figure 1.

**Figure 1.**
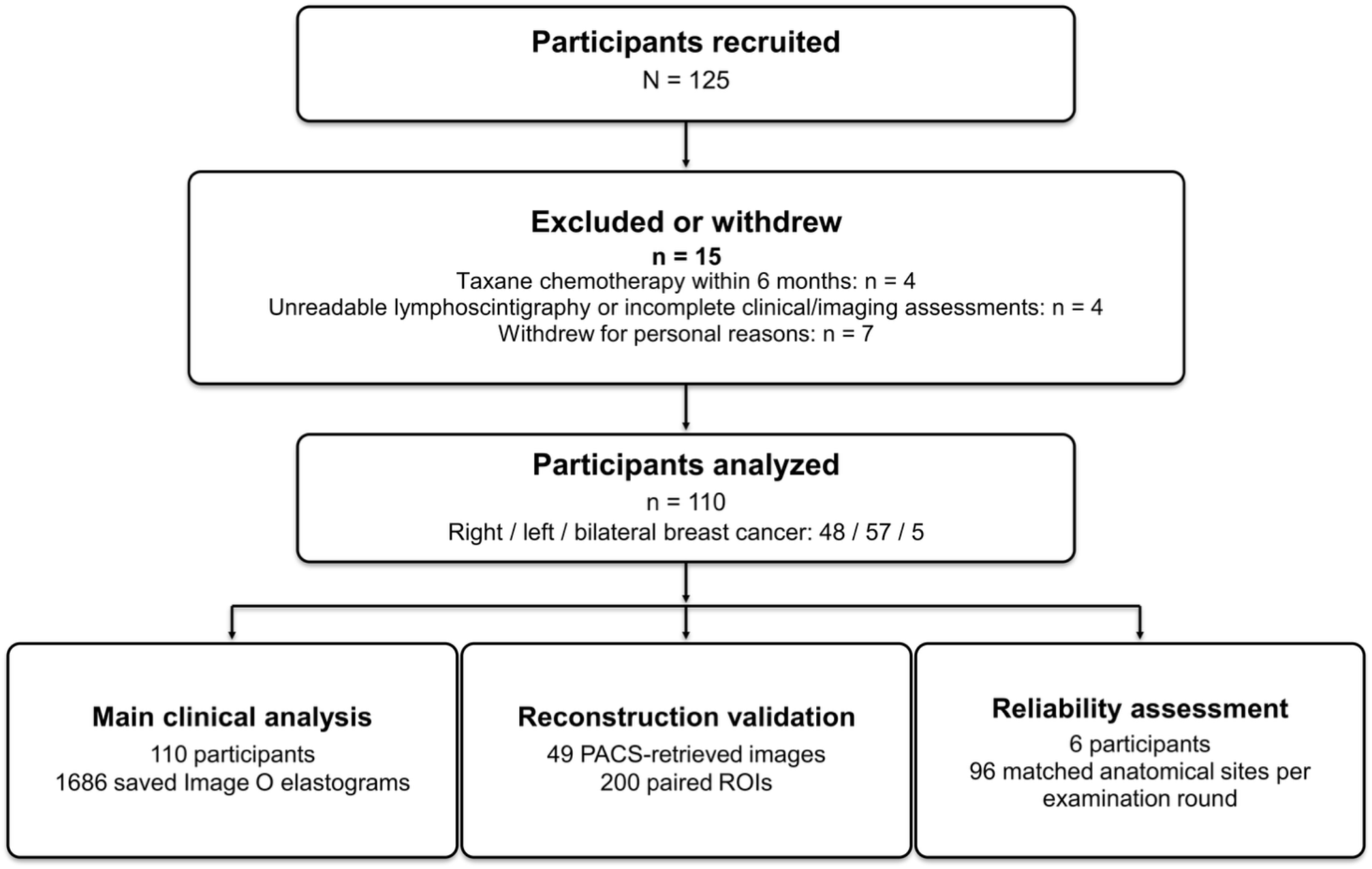
Participant flow and analytic subsets. The main clinical analysis included 110 participants and 1686 saved “Image O” elastograms; separate subsets were used for reconstruction validation (49 images, 200 paired ROIs) and reliability assessment (6 participants, 96 matched anatomical sites per examination round).

A separate clinical control group was formed from postoperative breast cancer participants without lymphedema-related symptoms. Because prior breast cancer treatment may alter ipsilateral lymphatic function even before overt lymphedema develops^19,20^, we used these postoperative participants as our clinical controls, rather than patients without a history of cancer. Lymphoscintigraphy and elastography were scheduled within 2 weeks on different days.

Enrollment data for all participants included breast cancer timing and stage, treatments, recurrence or metastasis status, lymphedema symptoms and onset, International Society of Lymphology (ISL) stage^21^, body mass index, and bilateral upper-limb circumferences.

Lymphoscintigraphy served as the imaging reference standard for identifying and grading lymphatic obstruction. Participants were classified by the Taiwan Lymphoscintigraphy Staging (TLS) system^5^ as having no, partial, or total obstruction. The primary diagnostic comparison was “no obstruction versus partial obstruction” (*no-versus-partial*) and the secondary comparisons were “no obstruction versus any obstruction” (*no-versus-any*) and “partial versus total obstruction” (*partial-versus-total*). Full TLS definitions and the protocol are provided in Supplement 2.

### Sonography acquisition and image sets

The images used in the present study were acquired as part of the HOE methodological study^18^. B-mode sonography and ARFI elastography were performed using an Acuson S2000 ultrasound system with Virtual Touch IQ and a 9L4 linear-array transducer (Siemens Medical Solutions, Mountain View, Calif; center frequency, 7.5 MHz; range, 4.0 – 9.0 MHz). Briefly, HOE uses an external apparatus to stabilize the ultrasound probe in a manner where only the coupling gel makes contact with the skin, thus minimizing applied pressure^18^ (Supplement 1). Imaging began 2.5 cm proximal to the wrist crease and extended toward the axilla along the palmar upper limb in transverse sections at 5-cm intervals. The wrist-crease region was avoided because the cutis and subcutaneous tissue are thin there and overlie superficial high-SWV structures such as tendons and ligaments. Depending on limb length, 6 to 8 positions were typically acquired per limb.

At each position, one 3-image set was obtained. In each set, “Image B” documented cutaneous and subcutaneous anatomy. ARFI elastography was then repeated until 3 consecutive elastograms showed a consistent superficial SWV distribution; the final unmarked image was saved as “Image O.” On the same underlying elastogram, five 1.0 × 1.0 mm ROIs were placed in the cutis and five in the subcutaneous tissue, and the marked image was saved as “Image M.” Thus, Image O served as the source for PDA reconstruction and visual HVA counting, while Image M provided machine-reported measurements for technical validation and the sparse manual-ROI comparator. PDA, visual HVA counting, and manual ROI sampling were therefore analytic frameworks applied to the same HOE-acquired elastograms.

An HVA was defined according to the HOE framework as a visually conspicuous focal subcutaneous region containing SWV values greater than 7 m/s^18^. All image files used for the main PDA analysis and the 200-ROI reconstruction validation were retrieved from the picture archiving and communication system (PACS) and retained in a non-destructive image format that preserved the color/SWV encoding. The three-image workflow is shown in Figure 2.

**Figure 2.**
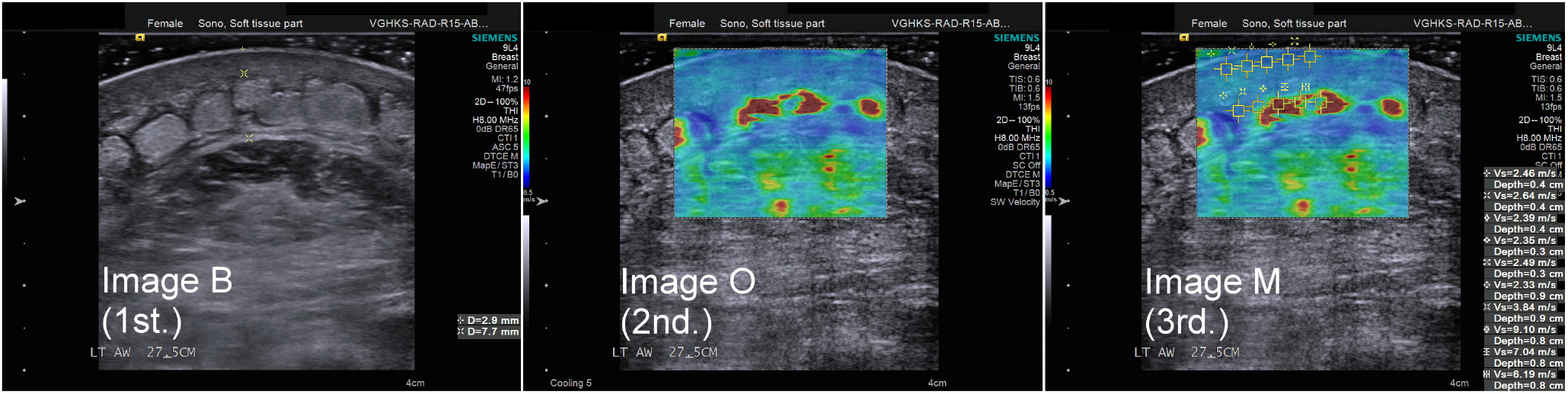
Representative HOE-acquired three-image set used for PDA. From left to right, Image B is the B-mode anatomic reference, Image O is the final consistent unmarked elastogram used for PDA reconstruction, and Image M is the marked machine-ROI image containing five cutaneous and five subcutaneous machine measurements used for validation and manual ROI comparison. The Image O panel illustrates the type of subcutaneous heterogeneity that HOE acquisition preserves and that PDA was designed to quantify.

A radiologist with 20 years of ultrasound experience supervised acquisition, confirmed anatomic targeting, and interpreted the images.

### Pixel-Level SWV Reconstruction Using Pixel Data Analysis

PDA was developed to recover quantitative pixel-level SWV information from saved composite elastograms, extending analysis from a small number of manually sampled ROIs to the full measurable color-encoded field. In the study-system images, SWV is displayed as a color overlay on the grayscale B-mode image. The red, green, and blue (RGB) values in a saved elastogram therefore contain contributions from both the SWV-encoded color signal and the underlying grayscale image and cannot be matched directly to the color bar (Figure 3).

**Figure 3.**
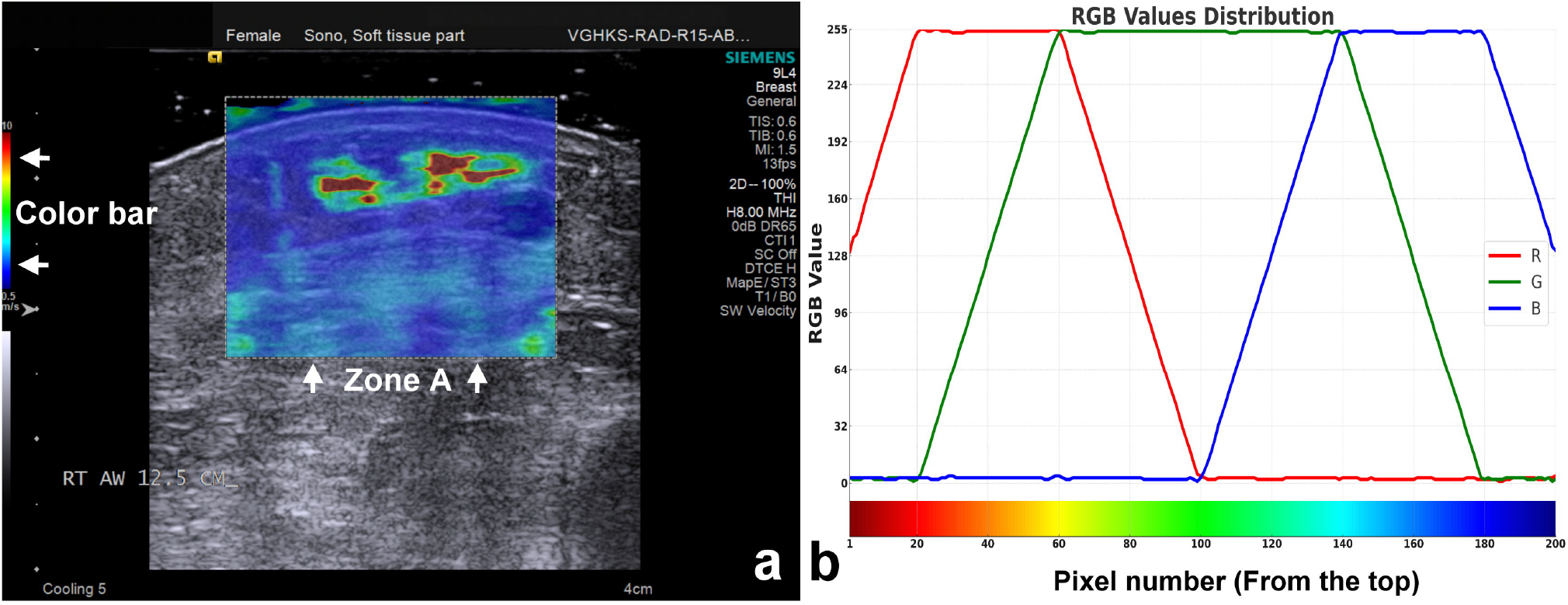
Study-system composite ARFI elastography image and color bar. a, Saved Image O showing the elastography box (Zone A) and study-system color bar. b, RGB progression along the color bar used to map displayed color to SWV.

Each observed pixel was modeled as a linear composite of the SWV-encoded color component and the underlying grayscale B-mode component:

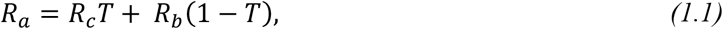

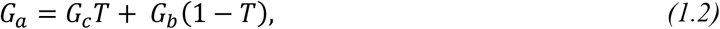

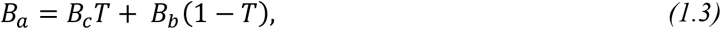

where (*R*_*a*_, *G*_*a*_, *B*_*a*_) were the RGB values in the saved elastogram; (*R*_*c*_, *G*_*c*_, *B*_*c*_) were the original SWV-encoding values; (*R*_*b*_, *G*_*b*_, *B*_*b*_) were the underlying B-mode values; and *T* was the transparency coefficient.

The decomposition was enabled by 2 properties of the study-system display. First, the underlying B-mode image was grayscale:

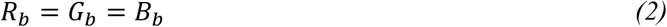

Second, at least one SWV-encoding channel – i.e., at least one of Rc, Gc, or Bc – was zero for every entry of the study-system color bar. Consequently, the minimum observed RGB channel represented the grayscale contribution to the composite pixel:

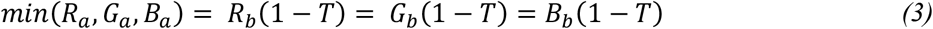

This minimum-channel relationship provided the basis for recovering the original SWV-encoded component.

The transparency coefficient was derived from the color-bar structure. When the observed green channel was dominant (i.e., *G*_*a*_= *max*(*R*_*a*_, *G*_*a*_, *B*_*a*_)), the corresponding SWV-encoding green channel was fixed at 255. Therefore:

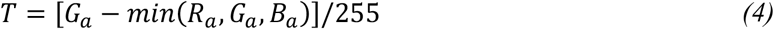

Under the acquisition and export settings used in this study, *T* was consistently 0.5. The original SWV-encoded RGB components could therefore be recovered directly from each composite pixel:

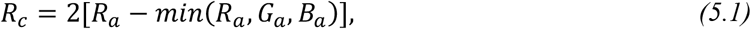

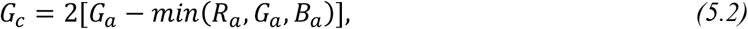

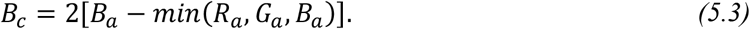

The recovered RGB values were then mapped to the calibrated study-system color bar to reconstruct physical SWV values. Although the displayed scale was labeled 0.5 – 10.0 m/s, image-based calibration showed that the stored color encoding extended from approximately 0.4 – 10.0 m/s. The conceptual reconstruction workflow is summarized in Table 1; system-specific mapping and validation details are provided in Supplement 3.

**Table 1.** Conceptual workflow for pixel-level SWV reconstruction using PDA.

| Step | Reconstruction principle | Output |
| --- | --- | --- |
| 1. Composite-image model | Saved elastogram pixels contain both grayscale B-mode and SWV-encoded color information. | Observed RGB values represent a composite rather than native SWV values. |
| 2. Grayscale identification | Because the underlying B-mode image is grayscale ($R_b = G_b = B_b$) and at least one SWV-encoding channel is zero for each color-bar entry, the minimum observed RGB channel identifies the grayscale contribution. | Grayscale and SWV-encoded components become separable. |
| 3. Transparency calibration | The study-system color-bar structure allows T to be solved from green-dominant pixels; T was consistently 0.5 under the study acquisition/export settings. | Original SWV-encoded RGB components can be recovered from saved composite pixels. |
| 4. Physical SWV reconstruction | Recovered RGB values are mapped to the calibrated study-system color bar. | A physical SWV value is reconstructed for each measurable color-encoded pixel. |
| 5. Whole-field quantification | PDA is applied to all measurable pixels within the operator-delineated cutaneous or subcutaneous layer. | Layer-specific pixel-level SWV datasets and distributional measures, including mean and variance. |

The technical validation used paired Image O and Image M files from the same underlying elastogram. Because the visible ROI border did not necessarily identify the system’s exact internal measurement coordinates after export, a small coordinate offset was allowed during matching. Spatial robustness of this procedure was evaluated by forced-displacement sensitivity analysis (Supplement 3).

Using the SWV Data Acquisition Applet (SDA app; Supplement 4), the operator delineated cutaneous and subcutaneous layers and PDA reconstructed the SWV of every measurable color-encoded pixel within each selected region. Outputs included pixel coordinates, physical SWV, and layer-specific distributional measures including mean, maximum, minimum, variance, and pixel count. This changed the sampling scale from a few manually selected ROIs to tens of thousands of reconstructed pixels within a tissue layer. Segmentation and export implementation are detailed in Supplement 4.

### Parameters and Data Pooling

Four layer-specific parameters were analyzed: cutaneous mean, cutaneous variance, subcutaneous mean, and subcutaneous variance. The prespecified PDA framework comprised 32 feature definitions generated from 4 layer-specific parameters, 4 spatial strategies, and 2 expressions. The 8 position-independent definitions analyzed individual imaging levels without cross-level pooling and were treated as image-level analyses. The remaining 24 definitions used global pooling across all available scan levels (2.5, 7.5, 12.5, 17.5, 22.5, 27.5, and, when available, 32.5 and 37.5 cm proximal to the wrist crease), joint-excluded pooling of the 7.5, 12.5, 17.5, 27.5, and, when available, 32.5 cm levels, or forearm pooling of the 7.5, 12.5, and 17.5 cm levels to generate participant-level metrics. For pooled analyses, reconstructed pixels from the relevant images were combined before the mean or variance was calculated. Direct values and affected-to-unaffected ratios were calculated within the same framework; participants with bilateral breast cancer were excluded from ratio analyses.

### Reliability assessment

Interobserver and intraobserver reliability were assessed in a dedicated 6-participant subset to reflect the cohort’s clinical spectrum: 1 postoperative participant without lymphedema and 1 participant from each of 5 ISL clinical strata^21^ (stage 0, stage I, early stage II, late stage II, and stage III). Each participant underwent 3 independently reacquired HOE-based ARFI examinations, with HOE setup, anatomical-site relocation, ARFI acquisition, tissue-layer delineation, and analysis repeated for each examination. Aa and Ab were performed by Examiner 1 on separate study days; B was performed by Examiner 2 on the same day as Ab. Each round included 16 matched anatomical sites per participant (8 per upper limb), giving 96 matched sites per round and 288 image instances across the 3 rounds. Intraobserver reliability compared Aa with Ab, and interobserver reliability compared Ab with B. Each intraclass correlation coefficient (ICC) comparison therefore used 96 site-level paired measurements. ICC(2,1) was estimated with a two-way random-effects, absolute-agreement, single-measurement model. Complete reliability results, site-target confidence interval (CI) details, and participant-level influence sensitivity analyses are provided in Supplement 5.

### Statistical analysis

The primary clinical comparison was no obstruction versus partial obstruction (“*no-versus-partial*”); secondary comparisons were no obstruction versus any obstruction (“*no-versus-any*”) and partial obstruction versus total obstruction (“*partial-versus-total*”).

The PDA framework comprised 32 prespecified feature definitions per task: 8 position-independent image-level definitions and 24 participant-level pooled candidates. The PDA headline metrics were selected by the highest observed area under the curve (AUC) among the 24 participant-level pooled candidates evaluated in their respective eligible screening cohorts, with bilateral cases retained in direct analyses and excluded from ratio analyses. For the framework comparison, the headline PDA metrics and the manual ROI metrics chosen in the original exploratory analysis were retained; the visual HVA metrics followed the related HOE visual-count framework instead^18^. Position-independent PDA analyses were evaluated separately at the image level to characterize the contribution of multilevel spatial integration and were not included in participant-level headline metric selection. Comparator AUCs were calculated in the same task-specific common complete-case cohorts: 93 participants for *no-versus-partial* obstruction (17 no, 76 partial), 105 for *no-versus-any* obstruction (17 no, 88 any), and 88 for *partial-versus-total* obstruction (76 partial, 12 total).

AUCs were reported with 95% CIs estimated using the DeLong variance method. Study-specific operating points were selected using the Youden index in the same cohorts, with sensitivity and specificity calculated at those points; these values are reported descriptively rather than as externally validated clinical cutoffs.

Reconstruction agreement was summarized with correlation, error metrics, and Bland-Altman limits of agreement. Reliability was summarized with ICC(2,1). Fixed-position analyses and the HVA-negative high-SWV exclusion sensitivity analysis are reported in Supplement 6. Internal performance of the fixed forearm-pooled subcutaneous variance ratio was evaluated using repeated stratified five-fold cross-validation with training-fold feature selection for exploratory alternative features. The results are reported in Supplement 7.

Conventional statistical analyses were performed using IBM SPSS Statistics for Windows, version 24.0 (IBM Corp, Armonk, NY, USA). Reconstruction agreement analyses, DeLong AUC confidence intervals, image-cluster bootstrap analyses, and repeated stratified five-fold cross-validation were performed in R (version 4.3.3; R Foundation for Statistical Computing, Vienna, Austria).

## Results

### Cohort and analytic subsets

Of 125 women recruited after breast cancer surgery, 15 were excluded or withdrew, leaving 110 participants (Figure 1). A total of 1686 Image O elastograms were available for image-level analyses. Validation used 49 images with 200 paired ROIs. Reliability included 6 participants, 96 matched anatomical sites per examination round, and 96 paired site-level measurements in each ICC comparison. Cohort characteristics and analysis subsets are summarized in Table 2.

**Table 2.** Cohort characteristics and analysis subsets.

| Characteristic | Value |
| --- | --- |
| Participants analyzed, n | 110 |
| No / partial / total obstruction, n | 18 / 80 / 12 |
| Age, years | 60.8 ± 10.9 |
| Body mass index, kg/m<sup>2</sup> | 25.2 ± 4.0 |
| Right-sided / left-sided / bilateral breast cancer, n | 48 / 57 / 5 |
| Total mastectomy / partial mastectomy, n | 63 / 47 |
| Axillary dissection / sentinel-node dissection, n | 61 / 49 |
| Radiotherapy yes / no, n | 56 / 54 |
| Time from surgery to lymphedema onset, months | 48.8 ± 70.4 |
| Time from onset to enrollment, months | 23.3 ± 35.0 |
| Saved elastography images included in image-level analyses, n | 1686 |

*Values are presented as mean ± standard deviation or counts. Time intervals were summarized for 92 participants, excluding the 18 clinical controls; months were calculated as the date difference in days divided by 30.*

#### PDA reconstruction validity

Across 200 paired ROIs, the full range of machine-reported SWV values was 1.17 – 9.97 m/s. PDA reconstruction closely matched machine measurements, with negligible error and no meaningful spatial offset (Table 3). These validation images were retrieved from PACS in the same non-destructive format used for the clinical PDA analysis.

**Table 3.** Validation summary metrics.

| Metric | Value |
| --- | --- |
| Paired ROIs analyzed, n | 200 |
| Machine-reference SWV, mean ± SD, m/s | 5.25 ± 2.59 |
| Machine-reference SWV range, m/s | 1.17 – 9.97 |
| Pearson correlation (r) | 0.999991 |
| Mean difference (PDA – machine), m/s | +0.0009 |
| Mean absolute error, m/s | 0.0075 |
| Root mean squared error, m/s | 0.0108 |
| 95% limits of agreement, m/s | -0.0202 to +0.0221 |
| Overall mean x-offset, pixels | 0.00 |
| Overall mean y-offset, pixels | -0.18 |

*PDA, pixel data analysis; ROI, region of interest; SWV, shear-wave velocity; SD, standard deviation.*

### Reliability: PDA versus manual ROI

Across 96 matched anatomical sites from independently reacquired examinations, PDA showed higher interobserver and intraobserver reliability than sparse manual ROI sampling. This was most notable for subcutaneous variance, where interobserver ICC was 0.709 for PDA versus 0.178 for manual ROI, and intraobserver ICC was 0.799 versus 0.560. PDA also showed higher subcutaneous mean-SWV ICCs than manual ROI. Skin mean-SWV ICC point estimates were close between methods, whereas the intraobserver skin-variance ICC was higher for PDA (Table 4).

**Table 4.** Reliability summary for PDA and manual ROI analysis.

| Method | Surface | Statistic | Interobserver ICC(2,1) (95% CI) | Intraobserver ICC(2,1) (95% CI) |
| --- | --- | --- | --- | --- |
| **PDA** | Skin | Mean | 0.755 (0.639 – 0.835) | 0.934 (0.902 – 0.955) |
|  | Subcutaneous | Mean | 0.783 (0.692 – 0.850) | 0.819 (0.741 – 0.876) |
|  | Skin | Variance | 0.285 (0.094 – 0.457) | 0.970 (0.956 – 0.980) |
|  | Subcutaneous | Variance | 0.709 (0.593 – 0.796) | 0.799 (0.712 – 0.861) |
| **Manual ROI** | Skin | Mean | 0.707 (0.576 – 0.800) | 0.929 (0.895 – 0.952) |
|  | Subcutaneous | Mean | 0.578 (0.429 – 0.697) | 0.674 (0.548 – 0.769) |
|  | Skin | Variance | 0.231 (0.036 – 0.409) | 0.584 (0.436 – 0.702) |
|  | Subcutaneous | Variance | 0.178 (-0.017 – 0.362) | 0.560 (0.405 – 0.683) |

*Interobserver and intraobserver reliability were estimated with ICC(2,1) from 96 matched anatomical sites; complete reliability results and site-target confidence intervals are provided in Supplement 5. PDA, pixel data analysis; ROI, region of interest; ICC, intraclass correlation coefficient; CI, confidence interval.*

### Primary comparison: no obstruction versus partial obstruction

Comparator results using a common complete-case cohort across all three frameworks within each task are summarized in Table 5. In the primary *no-versus-partial* comparison (n = 93), the forearm-pooled PDA subcutaneous variance ratio showed the highest observed AUC (0.858), followed by visual HVA counting (0.820) and manual ROI sampling (0.794; Table 5). The corresponding receiver operating characteristic (ROC) curves and PDA metric distributions are shown in Figure 4, and the study-specific operating points are also shown in Table 5.

**Table 5.**
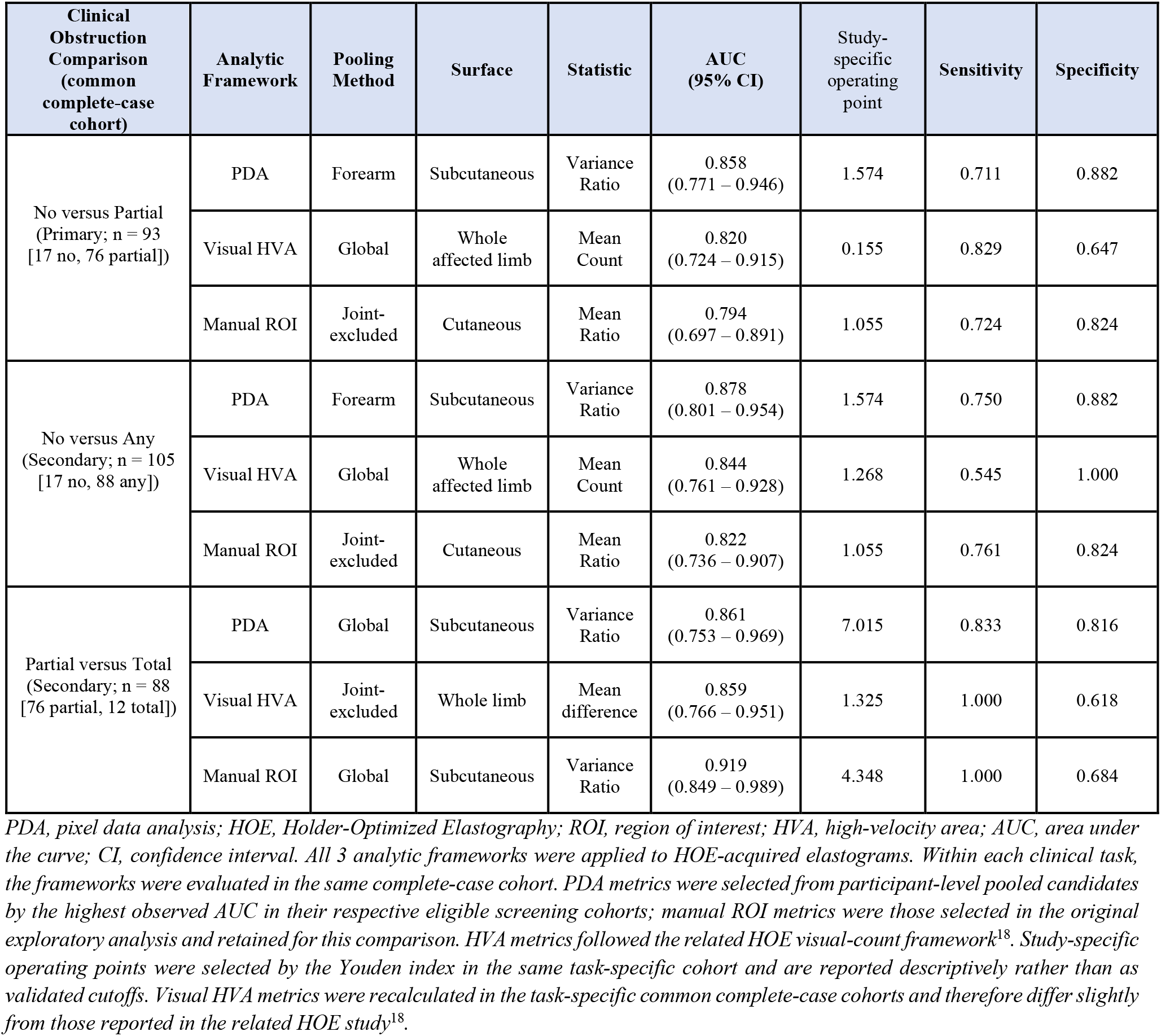
Comparator results by clinical task.

| Clinical Obstruction Comparison (common complete-case cohort) | Analytic Framework | Pooling Method | Surface | Statistic | AUC (95% CI) | Study-specific operating point | Sensitivity | Specificity |
| --- | --- | --- | --- | --- | --- | --- | --- | --- |
| No versus Partial (Primary; n = 93 [17 no, 76 partial]) | PDA | Forearm | Subcutaneous | Variance Ratio | 0.858 (0.771 – 0.946) | 1.574 | 0.711 | 0.882 |
|  | Visual HVA | Global | Whole affected limb | Mean Count | 0.820 (0.724 – 0.915) | 0.155 | 0.829 | 0.647 |
|  | Manual ROI | Joint-excluded | Cutaneous | Mean Ratio | 0.794 (0.697 – 0.891) | 1.055 | 0.724 | 0.824 |
| No versus Any (Secondary; n = 105 [17 no, 88 any]) | PDA | Forearm | Subcutaneous | Variance Ratio | 0.878 (0.801 – 0.954) | 1.574 | 0.750 | 0.882 |
|  | Visual HVA | Global | Whole affected limb | Mean Count | 0.844 (0.761 – 0.928) | 1.268 | 0.545 | 1.000 |
|  | Manual ROI | Joint-excluded | Cutaneous | Mean Ratio | 0.822 (0.736 – 0.907) | 1.055 | 0.761 | 0.824 |
| Partial versus Total (Secondary; n = 88 [76 partial, 12 total]) | PDA | Global | Subcutaneous | Variance Ratio | 0.861 (0.753 – 0.969) | 7.015 | 0.833 | 0.816 |
|  | Visual HVA | Joint-excluded | Whole limb | Mean difference | 0.859 (0.766 – 0.951) | 1.325 | 1.000 | 0.618 |
|  | Manual ROI | Global | Subcutaneous | Variance Ratio | 0.919 (0.849 – 0.989) | 4.348 | 1.000 | 0.684 |

PDA, pixel data analysis; HOE, Holder-Optimized Elastography; ROI, region of interest; HVA, high-velocity area; AUC, area under the curve; CI, confidence interval. All 3 analytic frameworks were applied to HOE-acquired elastograms. Within each clinical task, the frameworks were evaluated in the same complete-case cohort. PDA metrics were selected from participant-level pooled candidates by the highest observed AUC in their respective eligible screening cohorts; manual ROI metrics were those selected in the original exploratory analysis and retained for this comparison. HVA metrics followed the related HOE visual-count framework<sup>18</sup>. Study-specific operating points were selected by the Youden index in the same task-specific cohort and are reported descriptively rather than as validated cutoffs. Visual HVA metrics were recalculated in the task-specific common complete-case cohorts and therefore differ slightly from those reported in the related HOE study<sup>18</sup>.

**Figure 4.**
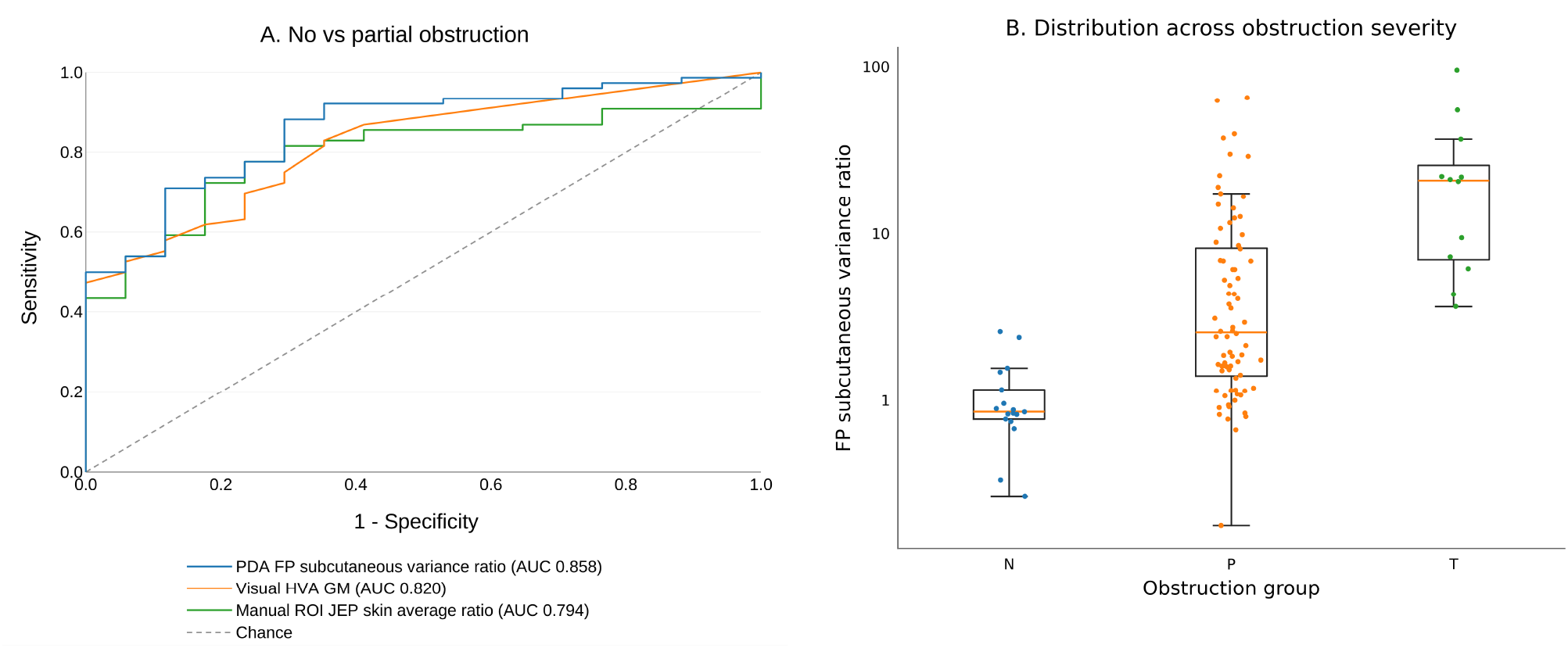
PDA in the primary comparison of distinguishing no obstruction from partial obstruction. A, ROC curves for no obstruction versus partial obstruction. B, Distribution of the forearm-pooled subcutaneous variance ratio across no obstruction (N), partial obstruction (P), and total obstruction (T). FP, forearm-pooled; GM, global mean; JEP, joint-excluded pooling; HVA, high-velocity area; ROI, region of interest; PDA, pixel data analysis; ROC, receiver operating characteristic; AUC, area under the curve.

### Secondary comparisons: no obstruction versus any obstruction and partial obstruction versus total obstruction

In the *no-versus-any* common cohort (n = 105), the leading PDA feature remained forearm-pooled subcutaneous variance ratio and yielded an AUC of 0.878, compared with 0.844 for visual HVA counting and 0.822 for manual ROI sampling.

In the *partial-versus-total* common cohort (n = 88), all 3 frameworks showed high discrimination. Manual ROI global-pooled subcutaneous variance ratio yielded the highest observed AUC (0.919), followed by global-pooled PDA subcutaneous variance ratio (0.861) and then visual HVA counting (0.859). Direct affected-limb subcutaneous variance, without contralateral normalization, remained informative across all three comparisons (AUC, 0.809 – 0.868; Supplement 6).

In separate image-level position-independent analyses, the same subcutaneous variance-ratio feature yielded lower AUCs than the corresponding participant-level pooled analyses for all three tasks (Supplement 6), supporting the contribution of multilevel spatial integration.

Across all 3 clinical comparisons, the leading PDA feature belonged to the same feature family, subcutaneous variance ratio; comparisons involving no obstruction favored forearm pooling, whereas *partial-versus-total* discrimination favored global pooling. Fixed-position analyses supported this spatial pattern, with the strongest *no-versus-partial* single-position discrimination at 17.5 cm and more spatially distributed *partial-versus-total* performance (Supplement 6).

This pooling strategy was also robust under two additional checks. In a regional visual-HVA-negative sensitivity analysis of 23 partial-obstruction participants, the forearm-pooled variance ratio remained discriminatory (AUC, 0.744) and retained an AUC of 0.696 after exclusion of all pixels above 3 m/s (Supplement 6). In repeated stratified five-fold cross-validations with training-fold selection of alternative subcutaneous features, the forearm-pooled subcutaneous variance ratio, which was held constant across folds, retained the highest mean held-out AUC among the evaluated subcutaneous feature-construction strategies (Supplement 7).

## Discussion

PDA reconstructed pixel-level subcutaneous SWV from single routinely saved elastograms of lymphedematous tissue with negligible bias, and this spatially comprehensive method showed better reproducibility of subcutaneous variance relative to sparse manual ROI sampling. In HOE-acquired images, whole-field PDA showed the highest observed AUC among the three analytic frameworks for the primary *no-versus-partial* and secondary *no-versus-any* comparisons, whereas all three frameworks showed high discrimination for *partial-versus-total* obstruction. Across all three PDA analyses, the leading feature belonged to the same subcutaneous-variance-ratio family, with forearm pooling favored in comparisons involving no obstruction and global pooling for *partial-versus-total* grading. These findings indicate that the magnitude, spatial extent, and intralimb distribution of subcutaneous SWV heterogeneity carry diagnostic information about lymphatic obstruction that scalar or amplitude-thresholded measures do not capture, provided that acquisition preserves this spatial information.

After system-specific calibration, PDA reconstructed physical SWV for every measurable pixel of a single routinely saved composite elastogram without requiring a paired B-mode image or native quantitative export for individual cases. Reconstruction showed negligible bias across 1.17 – 9.97 m/s (mean difference = +0.0009 m/s; 95% limits of agreement, -0.0202 to +0.0221 m/s). Across fully reacquired examinations, PDA yielded higher inter- and intraobserver ICCs for subcutaneous variance than sparse manual ROI sampling (0.709 vs 0.178 and 0.799 vs 0.560, respectively). Because each examination was fully reacquired, these ICCs reflect reproducibility of the complete acquisition-to-analysis workflow rather than repeated analysis of the same stored image. With non-destructive storage, no additional acquisition-time file is needed for later PDA analysis. Both validation and clinical images were retrieved from PACS through this pathway, so retrospective reanalysis requires no rescanning.

Other systems require additional system-specific color/SWV calibration and storage-pathway verification. Prior image-based SWE methods addressed parts of this offline-quantification problem. Zhong et al^22^ recovered pixel-level elasticity from a composite DICOM by subtracting the corresponding B-mode image, requiring that this information be retained at acquisition or export. Skerl et al^23^ enabled DICOM/JPG reanalysis without a paired image for each case but derived their adjusted color map using quantitative data export (QDE) native elasticity data, a research-only export unavailable on commercial systems; discrimination based on standard deviation remained lower than manual/QDE assessment. Lee et al^24^ used image-wide RGB color ratios rather than reconstructed physical SWV to evaluate muscle stiffness; this summarized color distribution without retaining pixel-wise physical SWV values and their spatial coordinates. After system-specific calibration and storage-pathway verification, PDA was able to reconstruct the physical values associated with each pixel while preserving the spatial data, without additional per-case preprocessing.

However, for PDA to quantify spatial information, acquisition must first preserve it. The HOE method used herein minimizes superficial loading. In a matched-site study, visually conspicuous subcutaneous HVAs were seen at 108 sites only with HOE and at none only with handheld acquisition^18^. The study also showed that HVA-associated abnormalities corresponded spatially to obstruction-associated lymphatic structures on MRI, D2-40 histopathology, and intraoperative indocyanine-green imaging. Therefore, HOE preserves the spatial SWE abnormality, and PDA quantifies its full measurable distribution, providing the basis for applying PDA to HOE-acquired elastograms. This concordant localization supports HVA and the broader SWE spatial abnormality as spatial markers of the location and distribution of obstructed lymphatic structures.

In evaluating these HOE-preserved abnormalities for diagnostic value, we previously found that counting these visually conspicuous HVAs captures a high-amplitude component of the HOE-preserved abnormality^18^, whereas in the present study, PDA quantifies the broader continuous SWV distribution. Less extensive obstruction may contain lower-amplitude or patchy abnormalities below the HVA threshold; advanced obstruction may have abundant HVAs in both partial and total categories. In the primary *no-versus-partial* comparison, PDA yielded the highest observed AUC (0.858), compared with 0.820 for HVA counting and 0.794 for manual ROI sampling.

In the secondary *no-versus-any* comparison, PDA likewise yielded the highest observed AUC (0.878), compared with 0.844 for HVA counting and 0.822 for manual ROI sampling. In *partial-versus-total* obstruction, manual ROI yielded the highest AUC (0.919), compared with 0.861 for PDA and 0.859 for visual HVA. Across the two secondary comparisons, the operating-point trade-offs differed among frameworks, whereas PDA showed a relatively balanced sensitivity-specificity profile while maintaining high AUCs.

All comparator analyses used the same HOE-acquired elastograms, so the differences primarily reflected analytic strategy rather than acquisition quality. Manual ROI sampling achieved a high AUC for partial versus total obstruction (0.919), but its subcutaneous-variance interobserver/intraobserver ICCs were 0.178/0.560, compared with 0.709/0.799 for PDA. Whole-field analysis therefore provided a more reproducible measure of heterogeneity and is particularly suited to abnormalities that are patchy or sensitive to sampling position.

Position-independent subcutaneous variance-ratio AUCs were lower than in the corresponding participant-level pooled analyses for all three comparisons, supporting the contribution of multilevel spatial integration. The spatial scale of the informative signal also changed with obstruction severity; comparisons involving no obstruction were forearm-predominant, with the strongest single-position *no-versus-partial* discrimination at 17.5 cm, whereas *partial-versus-total* discrimination was more spatially distributed and favored global pooling. This forearm-to-global shift indicates that both the magnitude and anatomical extent of subcutaneous heterogeneity contribute to obstruction grading, consistent with MRI evidence of spatially nonuniform earlier disease and broader involvement with increasing severity^16,17^.

This subcutaneous heterogeneity signal was also evident without contralateral normalization. In participant-level direct analyses, affected-limb subcutaneous variance remained informative across the clinical spectrum (AUC, 0.809 – 0.868). Concordant direct and ratio-based results indicate that disease-related information resides in the subcutaneous SWV distribution itself, while bilateral normalization additionally incorporates an internal reference.

Prior lymphedema SWE studies using scalar measurements captured only part of this soft-tissue spatial pattern. Chan et al^11^ found little change in mean cutaneous or subcutaneous SWV from partial to total obstruction; He et al^25^ found stage differences only in forearm cutis; and Bang et al^26^, despite using multiple ROIs to reduce sampling bias, detected SWE differences in clinical but not subclinical lymphedema and summarized the ROI measurements into a single site-level SWV estimate.

Lymphedema-specific ultrasound studies also support distribution-sensitive analysis. Sanderson et al^15^ described heterogeneous elastography maps despite inconsistent subcutaneous SWV differences, while Lee et al^27^ found that backscatter entropy discriminated lymphatic obstruction better than conventional amplitude-based measures. These observations indicate that mean measurements may incompletely represent disease-related variation within subcutaneous tissue.

### Limitations

This study had several limitations. It was conducted at a single center, used a cross-sectional design, and was limited to upper-extremity BCRL. Tissue-layer segmentation remained operator-guided. In addition, the total-obstruction subgroup was relatively small (n = 12), limiting the precision of estimates involving this category.

## Conclusion

PDA quantified HOE-preserved spatial information, yielding subcutaneous SWV heterogeneity as a reproducible diagnostic signal across the obstruction spectrum. PDA reconstructed this signal with negligible bias, and the same subcutaneous variance-ratio feature remained the leading PDA metric across all three clinical comparisons, with the informative spatial scale shifting from forearm pooling in comparisons involving no obstruction to global pooling for partial versus total obstruction. In 23 participants with partial obstruction and no visually detectable HVA at the three forearm levels, the forearm-pooled subcutaneous variance ratio remained discriminatory even after all pixels above 3 m/s were excluded (AUC, 0.696); the quantitative signal therefore extended beyond conspicuous high-velocity foci. The direct affected-limb analyses further indicate that PDA’s diagnostic value reflects the spatial distribution of subcutaneous heterogeneity preserved by HOE in addition to conspicuous HVA. Because reconstruction works from compatible PACS-retrieved elastograms, this quantitative analysis can be applied retrospectively to existing SWE archives without additional acquisition.

## Data Availability

All data produced in the present study are available upon reasonable request to the authors

