## Supplemental Documents for "Pixel-Level Quantification of Subcutaneous Shear-Wave Velocity Heterogeneity for Grading Lymphatic Obstruction in Breast Cancer-Related Lymphedema"

### Supplementary Material

*Citation numbers in this Supplementary Material correspond to the reference list in the main manuscript. The related HOE methodological report[18] is cited only where acquisition context is needed.*

#### Supplement 1. Holder-Optimized Elastography (HOE) Acquisition Components

All elastograms analyzed with PDA were acquired using the HOE framework described in the main manuscript and related methodological report[18]. The acquisition components relevant to the present image dataset are shown in S. Figure 1.

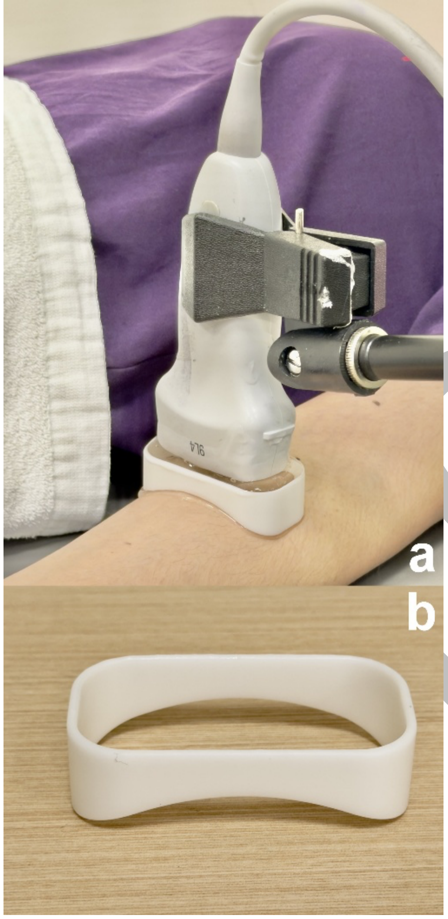


*S. Figure 1. HOE acquisition components used to generate the PDA dataset. a, Representative setup with the transducer externally stabilized above the limb and coupling gel as the skin interface. b, Gel holder used to maintain the coupling gel without direct probe-skin contact.*

Preprint

### Supplement 2. Lymphoscintigraphy Protocol and Taiwan Lymphoscintigraphy Staging (TLS) Classification

Planar lymphoscintigraphy and elastography of both upper extremities were performed within 2 weeks of each other. To avoid unnecessary radiation exposure to ultrasound operators and potential interference from residual tracer activity, lymphoscintigraphy and elastography were not performed on the same day. After subcutaneous injection of technetium-99m phytate into the first web space of each hand (2 mCi in 0.2 mL per hand; total dose, 4 mCi), static planar images were obtained at approximately 5 – 10 minutes and again at 2 – 3 hours after injection. Final interpretation was determined by consensus between 2 nuclear medicine physicians.

TLS was selected as the imaging reference standard because it provides an established imaging-based categorization of lymphatic obstruction severity in this clinical context[5]. For the present study, TLS L-0 was treated as no obstruction, P-1 to P-3 as partial obstruction, and T-4 to T-6 as total obstruction.

### Supplement 3. PDA Implementation and Reconstruction Validation

*3.1 Study-system calibration and separation verification*

The mathematical decomposition of the saved composite elastogram is described in the main Methods. For implementation, the elastography box (Zone A) measured 330 × 420 pixels, and the study-system color bar shown Figure 3 of the main text was 200 pixels long. Although the displayed scale was labeled 0.5 – 10.0 m/s, image-based calibration showed that the stored color encoding extended from approximately 0.4 to 10.0 m/s. The exact recovered-RGB-to-SWV mapping rules are provided in S. Table 1.

A representative separation result is shown in S. Figure 2a-f. The recovered grayscale component closely reproduces the paired B-mode appearance within Zone A, providing a visual check of the separation step. Panel g provides a pixel-level example of the compositing problem: two visually similar adjacent pixels can have different observed RGB values because their underlying grayscale contributions differ.

Preprint

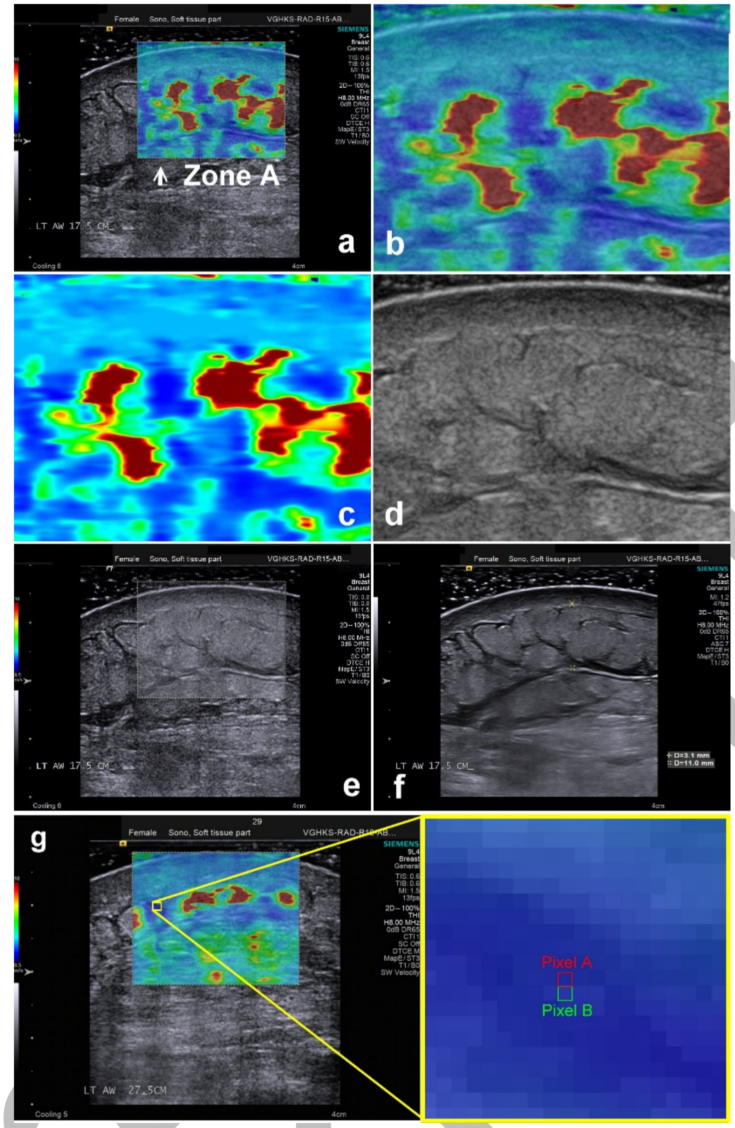


*S. Figure 2. Verification of grayscale and SWV-encoded color separation and the effect of grayscale compositing. a, Original Image O. b, Extracted Zone A. c, Recovered color-encoded SWV component. d, Recovered grayscale component. e, Reconstructed grayscale component overlaid back onto Zone A. f, Corresponding Image B at the same location. g, Pixel-level example showing why direct RGB-to-SWV matching is not valid: two visually similar adjacent pixels, A and B, had observed RGB values of (30, 43, 156) and (41, 52, 167), respectively, because their underlying grayscale B-mode contributions differed.*

#### *3.2 Pixel-by-pixel SWV reconstruction rules*

After recovery of the SWV-encoded RGB values, each pixel was assigned an SWV value by interpolation against the calibrated study-system color bar. During method refinement using 10 ARFI elastography images, minor RGB deviations introduced during display capture and image saving were typically within ±3 channel units and produced minimal SWV error. The final mapping therefore incorporated this tolerance and the empirically observed 0.4-m/s lower bound.

The reconstruction procedure used in the present study was as follows:

Step 1: Retrieve the original RGB values (Ra, Ga, Ba) of any pixel in Zone A.

Step 2: Subtract the smallest value among Ra, Ga, and Ba from each component, then multiply the results by 2 to obtain (Rc, Gc, Bc).

Step 3: Since RGB values are integers in the range of 0 – 255, reset any values in (Rc, Gc, Bc) exceeding 255 to 255, yielding values (r, g, b).

Step 4: Apply the clipped recovered RGB values (r, g, b) to the rule-based mapping scheme summarized in S. Table 1. This returns v, which represents the reconstructed SWV value for the pixel.

**S. Table 1. Rule-based mapping summary for pixel-level SWV reconstruction from recovered RGB values**

| Dominant recovered channel condition | Additional condition | Reconstruction rule for v (m/s) |
|---|---|---|
| b >= r and b >= g | b < 128 | v = 0.4 |
| b >= r and b >= g | $\lvert r - g\rvert$ <= 2 | v = (b - 127) * 0.0075 + 0.4 |
| b >= r and b >= g | $\lvert r - g\rvert$ > 2 and g < r | v = (b - 127) * 0.00735 + 0.4 |
| b >= r and b >= g | $\lvert r - g\rvert$ > 2 and g > r | v = g * 0.00735 + 1.408 |
| g >= r and g >= b | $\lvert b - r\rvert$ > 2 and b > r | v = (255 - b) * 0.00735 + 3.328 |
| g >= r and g >= b | $\lvert b - r\rvert$ > 2 and r > b | v = r * 0.00735 + 5.248 |
| g >= r and g >= b | $\lvert b - r\rvert$ <= 2 | v = 5.248 |
| r >= g and r >= b | r <= 130 | v = 10 |
| r >= g and r >= b | $\lvert g - b\rvert$ > 2 and g > b | v = (255 - g) * 0.00735 + 7.168 |
| r >= g and r >= b | $\lvert g - b\rvert$ > 2 and b > g | v = (255 - r) * 0.00735 + 9.088 |
| r >= g and r >= b | $\lvert g - b\rvert$ <= 2 | v = (255 - r) * 0.0073 + 9.088 |

*Notes: r, g, and b are the clipped recovered RGB values obtained after grayscale separation and rescaling in Step 3. The threshold for nearly equal channels was defined as an absolute channel difference of 2 or less. When ties occurred, branch assignment followed the implementation order used in the study, namely blue-dominant first, then green-dominant, and finally red-dominant. This table summarizes the rule-based mapping used to reconstruct pixel-level SWV values from recovered RGB data.*

Preprint

*3.3 Coordinate-offset handling*

Coordinate-offset handling is illustrated in S. Figure 3.

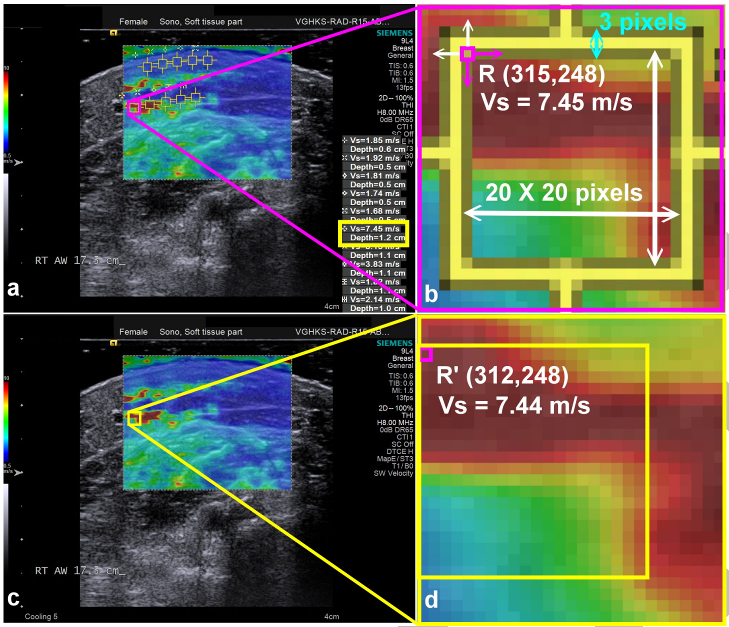


*S. Figure 3. Coordinate-offset handling for ROI-based validation. a, Machine ROI displayed on Image M. b, The same ROI represented as a 20 × 20-pixel region with a 3-pixel border. c, Candidate regions searched within ±3 pixels in both x- and y-directions in paired Image O. d, Closest-matching reconstructed region identified for validation.*

Each machine ROI displayed on Image M represented a 20 × 20-pixel measurement region, while the visible yellow border was 3 pixels wide. Because the system's internal measurement coordinate is not exposed after image export, the displayed border may not coincide exactly with the internal measurement region.

Validation therefore evaluated all candidate 20 × 20-pixel regions whose top-left coordinates fell within ±3 pixels in both x- and y-directions around the displayed ROI coordinate and selected the region with the closest reconstructed mean SWV for paired comparison.

To test the spatial specificity of coordinate matching, deliberate displacement from the PDA-selected ROI origin was evaluated across the same 200 validation points from 49 images. Local gradient sensitivity strongly predicted the increase in absolute error produced by a 10-pixel displacement (Spearman $\rho = 0.9514$; image-cluster bootstrap 95% CI, 0.9282 – 0.9648), supporting the spatial specificity of the coordinate-matching procedure.

*3.4 Validation sampling and interval performance*

Validation used paired Image O and Image M files from the same underlying elastogram. This section provides the sampling and range-specific details that supplement the aggregate validation results reported in the main manuscript.

After excluding the 10 images used during derivation and refinement, 49 ARFI elastography image pairs were randomly selected for validation and retrieved from PACS in the same non-destructive format used for the clinical

Preprint

PDA analysis. Within these images, 200 validation ROIs were sampled across machine-reported SWV strata to span the displayed range. ROIs were selected when the ROI and surrounding region showed relatively uniform color tone to minimize local heterogeneity as a source of coordinate-matching error.

At least 20 ROI measurements were included within each 1-m/s interval from 1 to 10 m/s. The resulting machine-reference values were 5.25 ± 2.59 m/s (range, 1.17 – 9.97 m/s).

Across the nine populated 1-m/s strata, absolute bias did not exceed 0.0043 m/s, and mean absolute error ranged from 0.0025 to 0.0119 m/s.

Aggregate agreement metrics are reported in main-text Table 3. Interval-specific errors are shown in S. Table 2, and the full scatter and Bland-Altman plots are shown in S. Figure 4.

**S. Table 2. Validation error summary by machine SWV interval**

| Machine SWV bin | n | Machine mean | PDA mean | Bias | SD of difference | MAE | RMSE |
|---|---|---|---|---|---|---|---|
| 0-1 | 0 | — | — | — | — | — | — |
| 1-2 | 25 | 1.670 | 1.668 | -0.0011 | 0.0030 | 0.0028 | 0.0032 |
| 2-3 | 25 | 2.519 | 2.519 | -0.0002 | 0.0031 | 0.0025 | 0.0031 |
| 3-4 | 25 | 3.423 | 3.425 | +0.0019 | 0.0058 | 0.0046 | 0.0060 |
| 4-5 | 25 | 4.453 | 4.452 | -0.0010 | 0.0137 | 0.0090 | 0.0135 |
| 5-6 | 20 | 5.532 | 5.531 | -0.0011 | 0.0152 | 0.0118 | 0.0149 |
| 6-7 | 20 | 6.396 | 6.400 | +0.0043 | 0.0115 | 0.0093 | 0.0120 |
| 7-8 | 20 | 7.405 | 7.408 | +0.0028 | 0.0152 | 0.0119 | 0.0151 |
| 8-9 | 20 | 8.549 | 8.549 | +0.0009 | 0.0146 | 0.0112 | 0.0143 |
| 9-10 | 20 | 9.572 | 9.575 | +0.0030 | 0.0082 | 0.0068 | 0.0085 |

*Machine bins are based on the machine-reported ROI value. Bias is PDA minus machine. All error metrics are reported in m/s.*

Preprint

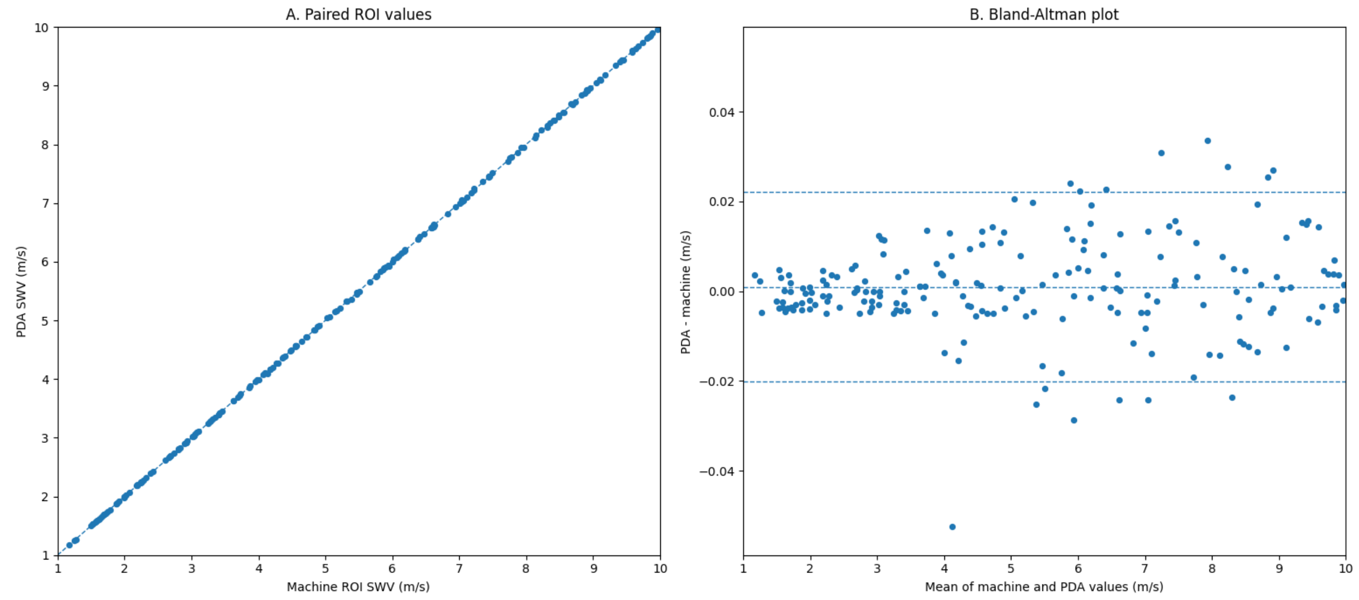


*S. Figure 4. Full validation display. A, Scatter plot comparing PDA-reconstructed SWV values with machine ROI values. B, Bland-Altman plot of paired values.*

### Supplement 4. SWV Data Acquisition Applet (SDA app): Segmentation and Export Implementation

To standardize application of PDA to saved Image O elastograms, we developed the SWV Data Acquisition Applet (SDA app) in HTML and Java for browser-based use. The app processes non-destructively preserved Image O files, including PACS-retrieved files; the operator interface is shown in S. Figure 5.

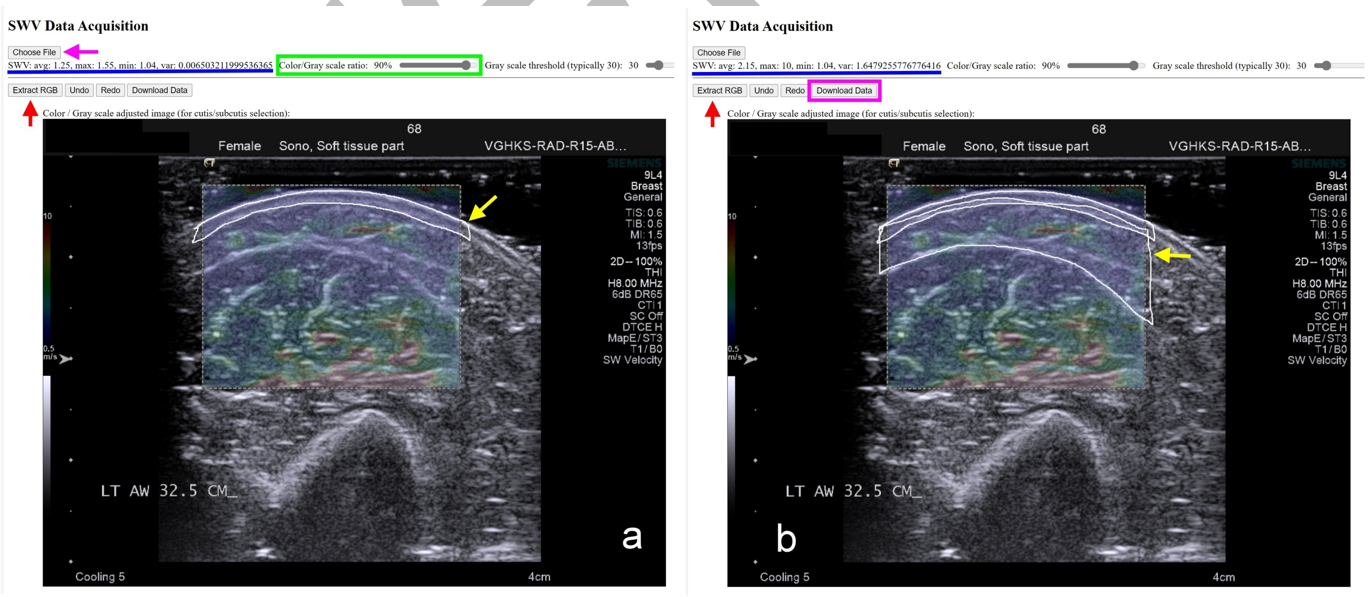


*S. Figure 5. SDA app interface for operator-guided segmentation. a, File loading and cutaneous-region delineation. b, Subsequent subcutaneous-region delineation after exclusion of the cutaneous region. The app reconstructs pixel-level SWV and allows export of the selected-region data.*

The operator can adjust the displayed color/grayscale balance to improve visualization of tissue-layer boundaries. The cutaneous region is delineated first and all measurable color pixels within the selected region are reconstructed; the subcutaneous region is then delineated after exclusion of the cutaneous region.

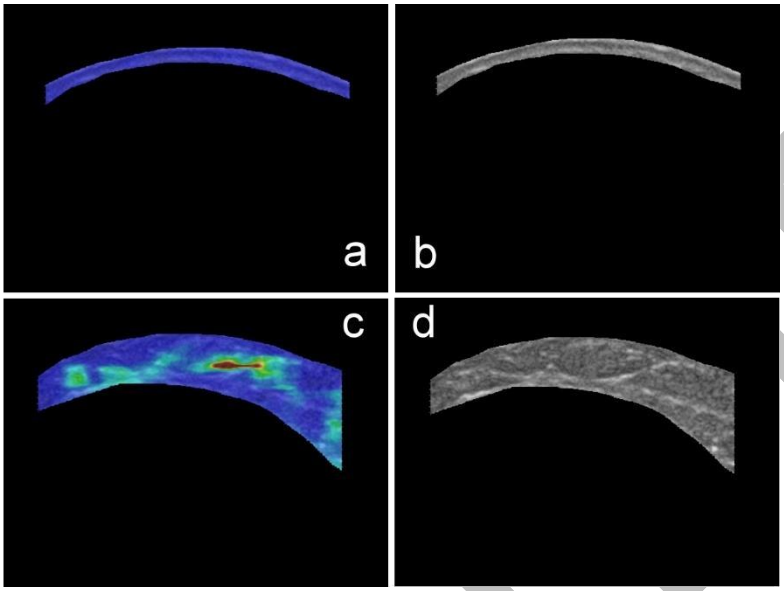


*S. Figure 6. Layer-selection verification displays generated by the SDA app. a and c, Selected cutaneous and subcutaneous regions, respectively. b and d, Corresponding recovered grayscale components used to verify that the outlined regions match the intended anatomy.*

After each delineation, the selected color-containing region and corresponding recovered grayscale component are displayed for anatomical verification (S. Figure 6). Exported outputs include the reconstructed SWV of each included pixel, pixel coordinates, and summary measures including mean, maximum, minimum, variance, and pixel count. In one representative subcutaneous segmentation, this workflow yielded 33,738 reconstructed pixel values.

The present implementation is system-specific because reconstruction depends on the calibrated study-system color encoding and transparency relationship. Application to another platform requires recalibration and validation of that platform's image-export and storage pathway.

In this study, the same physician who performed shear-wave elastography operated the SDA app to delineate cutaneous and subcutaneous regions and export the reconstructed pixel-level datasets. All clinical and technical-validation images were PACS-retrieved in a non-destructive format that preserved the composite grayscale/color encoding.

### Supplement 5. Reliability and Reproducibility

Complete reliability results are provided in S. Table 3. Aa, Ab, and B were fully reacquired examinations; each repeated HOE setup, anatomical-site relocation, ARFI acquisition, tissue-layer delineation, and analysis. Aa and Ab were performed by examiner 1 on separate study days; B was performed by examiner 2 on the same day as Ab. Interobserver reliability compared Ab with B and intraobserver reliability compared Aa with Ab. Each of the 96 participant-specific anatomical sites was treated as a distinct reliability target. ICC(2,1) was estimated with a two-



Preprint

way random-effects, absolute-agreement, single-measurement model, with conventional model-based 95% CIs for the 96 site targets.

**S. Table 3. Full ICC results for manual ROI and PDA**

| Method | Tissue layer | Statistic | Comparison | ICC(2,1) (95% CI) | Matched sites, n |
|---|---|---|---|---|---|
| PDA | Skin | Mean | Interobserver | 0.755 (0.639 – 0.835) | 96 |
| PDA | Skin | Mean | Intraobserver | 0.934 (0.902 – 0.955) | 96 |
| PDA | Skin | Variance | Interobserver | 0.285 (0.094 – 0.457) | 96 |
| PDA | Skin | Variance | Intraobserver | 0.970 (0.956 – 0.980) | 96 |
| PDA | Subcutaneous | Mean | Interobserver | 0.783 (0.692 – 0.850) | 96 |
| PDA | Subcutaneous | Mean | Intraobserver | 0.819 (0.741 – 0.876) | 96 |
| PDA | Subcutaneous | Variance | Interobserver | 0.709 (0.593 – 0.796) | 96 |
| PDA | Subcutaneous | Variance | Intraobserver | 0.799 (0.712 – 0.861) | 96 |
| Manual ROI | Skin | Mean | Interobserver | 0.707 (0.576 – 0.800) | 96 |
| Manual ROI | Skin | Mean | Intraobserver | 0.929 (0.895 – 0.952) | 96 |
| Manual ROI | Skin | Variance | Interobserver | 0.231 (0.036 – 0.409) | 96 |
| Manual ROI | Skin | Variance | Intraobserver | 0.584 (0.436 – 0.702) | 96 |
| Manual ROI | Subcutaneous | Mean | Interobserver | 0.578 (0.429 – 0.697) | 96 |
| Manual ROI | Subcutaneous | Mean | Intraobserver | 0.674 (0.548 – 0.769) | 96 |
| Manual ROI | Subcutaneous | Variance | Interobserver | 0.178 (-0.017 – 0.362) | 96 |
| Manual ROI | Subcutaneous | Variance | Intraobserver | 0.560 (0.405 – 0.683) | 96 |

*To assess participant-level influence, the pooled subcutaneous-variance ICC analysis was repeated six times, each time omitting one participant and all anatomical sites contributed by that participant. The PDA-over-manual pattern was preserved in all six analyses: interobserver ICCs ranged from 0.560 to 0.728 for PDA and 0.034 to 0.243 for manual ROI sampling, while intraobserver ICCs ranged from 0.530 to 0.821 and 0.275 to 0.587, respectively.*

### Supplement 6. Spatial Support and HVA-Negative Sensitivity Analysis

#### *6.1 Fixed-position spatial support*

Fixed-position analyses are shown in S. Figure 7. *No-versus-partial* discrimination was concentrated at forearm levels, with the highest single-position AUC at 17.5 cm (0.845; 95% CI, 0.768 – 0.923), whereas *partial-versus-total* performance was more spatially dispersed.

In separate position-independent image-level analyses, subcutaneous variance ratio yielded AUC point estimates of 0.715, 0.741, and 0.738 for *no-versus-partial*, *no-versus-any*, and *partial-versus-total* obstruction, respectively. The corresponding numbers of paired-position observations were 710 (93 participants), 803 (105 participants), and 669 (88 participants).

In participant-level direct analyses without contralateral normalization, subcutaneous variance yielded AUCs of 0.845 (n = 98) for *no-versus-partial* obstruction with forearm pooling, 0.868 (n = 110) for *no-versus-any* obstruction with joint-excluded pooling, and 0.809 (n = 92) for *partial-versus-total* obstruction with joint-excluded pooling.

Preprint

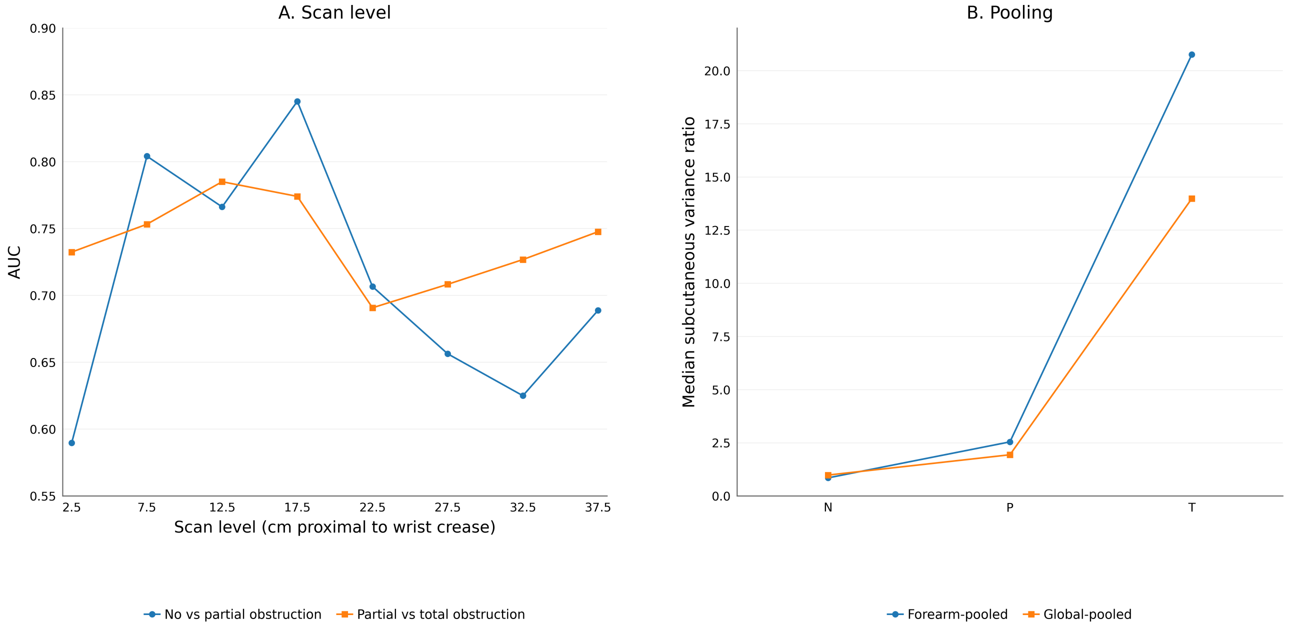


*S. Figure 7. Fixed-position analyses supporting forearm and global pooling. A, AUCs for single-position subcutaneous variance ratio across fixed scan levels (2.5 – 37.5 cm proximal to the wrist crease) in the no-versus-partial and partial-versus-total comparisons. B, Median forearm-pooled and global-pooled subcutaneous variance ratios across no obstruction (N), partial obstruction (P), and total obstruction (T). The 32.5- and 37.5-cm estimates are available-case because these proximal levels were not available in every participant. Ratio analyses excluded five bilateral cases. Panel B included 105 participants (N = 17, P = 76, T = 12). For panel A, effective n for no-versus-partial and partial-versus-total was 93 and 88 at 2.5 – 27.5 cm, 89 and 84 at 32.5 cm, and 63 and 57 at 37.5 cm, respectively.*

### *6.2 HVA-negative high-SWV exclusion sensitivity analysis*

To determine whether the whole-field variance signal was confined to conspicuous high-velocity foci, a regional visual-negative sensitivity analysis included 23 ratio-eligible partial-obstruction participants with no visually detected HVA at the three primary forearm sites (7.5, 12.5, and 17.5 cm), compared with 17 ratio-eligible no-obstruction participants. Forearm-pooled subcutaneous variance ratio was recalculated after progressively excluding pixels above prespecified SWV caps. Discrimination persisted after high-SWV exclusion, with AUC 0.696 even after all pixels above 3 m/s were removed (S. Table 4).

**S. Table 4. Regional visual-HVA-negative high-SWV exclusion sensitivity analysis**

| High-SWV exclusion condition | AUC |
|---|---|
| Unrestricted SWV | 0.744 |
| Exclude >6 m/s | 0.742 |
| Exclude >5 m/s | 0.714 |
| Exclude >4 m/s | 0.714 |
| Exclude >3 m/s | 0.696 |

*All cap conditions used the same participants and were treated as nested sensitivity analyses.*

### Supplement 7. Cross-Validation Performance Comparison

For the primary *no-versus-partial* task, repeated stratified five-fold cross-validation (50 repeats) compared the locked forearm-pooled subcutaneous variance ratio with an alternative subcutaneous feature selected within each training fold and with a model combining the two. The locked feature had been selected in the full-cohort analysis and was held fixed across all splits. The locked feature retained the highest mean held-out AUC among the evaluated strategies (S. Table 5).

**S. Table 5. Repeated stratified five-fold cross-validation performance comparison**

| Strategy | Mean held-out AUC |
|---|---|
| Locked variance | 0.861 |
| Training-fold-selected alternative | 0.777 |
| Locked + training-fold-selected alternative | 0.811 |

*Locked variance denotes the forearm-pooled subcutaneous variance-ratio feature selected in the full-cohort analysis and held fixed across folds. Training-fold-selected alternative denotes the best alternative feature selected exclusively within each training fold. Locked + training-fold-selected alternative denotes a model including the locked variance feature together with the training-fold-selected alternative. Values are mean held-out AUCs across 50 repetitions of stratified five-fold cross-validation in the primary no-versus-partial cohort (n = 93). The mean held-out AUC is a cross-validation performance estimate and is distinct from the full-cohort observed AUC reported in main-text Table 5.*

Preprint
